# Protocol for a prospective cohort study for the assessment of sarcopenia in gestational diabetes: The SiGnal-D study

**DOI:** 10.1101/2025.05.02.25326744

**Authors:** Angela Dardano, Giuseppe Daniele, Christian Göbl, Andrea Tura

## Abstract

**Background:** Sarcopenia is characterized by loss of muscle mass and strength. Although aging is the most likely sarcopenia risk factor, sarcopenia is frequent even in non-elderly people. Type 2 diabetes (T2D) is a risk factor for sarcopenia, as T2D shares with sarcopenia several etiological factors. On the other side, gestational diabetes mellitus (GDM) is characterized by metabolic alterations similar to those of T2D, although its distinctive trait is insulin resistance (present even in physiologic pregnancies). Hence, GDM presents two major risk factors for sarcopenia, i.e., dysglycemia and insulin resistance. Moreover, the number of pregnancies above 40 years is increasing, therefore in an age range where sarcopenia prevalence is already not negligible. However, there is lack of data about sarcopenia prevalence in GDM and about the impact on pregnancy outcomes. Thus, we plan evaluating sarcopenia prevalence in women with GDM (and in non-GDM pregnant women), identifying risk factors, and determining the impact on delivery as well as on maternal and fetal outcomes.

**Methods:** We will recruit 100 GDM and 100 non-GDM women. All women will undergo oral glucose tolerance test within week 24-28 for possible GDM diagnosis. Muscle/physical performance tests will be performed at week 28-32 for possible diagnosis of sarcopenia/presarcopenia. Cognitive function will also be assessed. For all women we will collect information regarding pregnancy progression, along with any complication. Collected data will be analyzed according to the study main objectives: i) determining the prevalence of sarcopenia/presarcopenia in pregnancy with and without GDM, ii) identifying factors associated with sarcopenia risk, iii) determining the impact of sarcopenia/presarcopenia on pregnancy outcomes, iv) exploring the relationship between sarcopenia and cognitive function.

**Discussion:** The present study will provide information on sarcopenia/presarcopenia prevalence in GDM and, possibly, in pregnancy uncomplicated by dysglycemia. Furthermore, the study will provide knowledge on the main factors associated to sarcopenia/presarcopenia in GDM/pregnancy. The identification of such factors will be relevant for an initial guidance for treatments that may prevent sarcopenia in GDM/pregnant women. This will become of even greater interest if sarcopenia/presarcopenia shows an impact on pregnancy outcomes, especially in GDM women.

**Trial registration:** ClinicalTrials.gov Identifier: NCT06876090; Registration Date: 2025-03-14

## Background

Sarcopenia is a syndrome characterized by loss of skeletal muscle mass and strength, with risk of adverse outcomes such as poor quality of life, physical disability and even death [1]. Recently, associations between sarcopenia and impaired cognitive function have been described as well [2]. Reduced muscle mass, along with low muscle strength or low physical performance, are required for the diagnosis of sarcopenia [3, 4]. The concomitance of the three factors characterizes severe sarcopenia. On the other hand, isolated muscle mass loss is better defined as presarcopenia [5]. With regard to the underlying mechanisms, it is possible to distinguish age-related sarcopenia (i.e., primary sarcopenia) and secondary sarcopenia, when other factors such as malnutrition (inadequate dietary intake of energy and/or proteins), insufficient physical activity, and presence of other diseases or disorders play a pathogenetic role [6]. Comorbidities favoring sarcopenia are gastrointestinal diseases (malabsorption), organ failure (e.g., heart, kidneys, lungs, brain and liver), inflammatory diseases, cancer, neurodegenerative diseases and metabolic/endocrine disorders or diseases (insulin resistance, type 2 diabetes, obesity). In addition, coexistence of sarcopenia and obesity identifies a specific sarcopenic phenotype, also known as sarcopenic obesity. Also, cachexia is often associated with sarcopenia [7], and pharmacological agents (such as corticosteroids) can also increase sarcopenia risk [8]. Of note, sarcopenia may be prevented [9], but, once manifest, remission is not common. However, pharmacologic intervention and adequate physical exercise can mitigate the clinical picture [10].

In recent years, prevalence of sarcopenia has been reported to increase. A recent review/meta-analysis analyzed 151 studies for a total of 692,056 subjects [11]. Sarcopenia prevalence ranged from 10% to 27% in 60+ years and, surprisingly, from 8% up to 36% in individuals below 60 years. This meta-analysis [11] showed that although aging is the most likely risk factor for sarcopenia, this condition can be frequent even in non-elderly people. This is clearly shown in studies where assessment of sarcopenia prevalence was reported for different age categories [12–14]. Pongchaiyakul et al. [12] (832 subjects studied) reported sarcopenia prevalence from 11% to 36% until 49 years. Bae et al. [13] (17,968 subjects), in the 20-39 years group, found sarcopenia prevalence at 19.2%. A lower, yet remarkable 9% prevalence has been reported by Cho et al. [14], in the below 50 years group (8,092 subjects, average age 38 years).

As mentioned above, type 2 diabetes (T2D) is a risk factor for sarcopenia. This is most likely due to the fact that T2D shares with sarcopenia several etiological factors, such as inflammation, oxidative stress, insulin resistance, cardiovascular comorbidities, low physical activity, malnutrition, and aging [15, 16]. On the other side, gestational diabetes mellitus (GDM) is characterized by metabolic alterations similar to those of T2D (mainly, insulin resistance and beta-cell dysfunction) [17], although its most distinctive trait is insulin resistance [18]. In fact, due to the hormonal changes during pregnancy, a certain degree of insulin resistance develops even in physiologic pregnancies [19]. Hence, GDM presents at least two major risk factors for sarcopenia, i.e., dysglycemia and severe insulin resistance. Moreover, the number of pregnancies above 40 years is increasing, therefore occurring in an age range in which the prevalence of sarcopenia is already sizeable [12–14]. Nevertheless, there is lack of data regarding the prevalence of sarcopenia in women with gestational diabetes mellitus (GDM) and its potential effects on maternal, fetal, and neonatal health. Thus, we plan to carefully evaluate sarcopenia prevalence in women with GDM (as well as in pregnant women without GDM), identify risk factors, and determine the potential impact on the pregnancy outcomes.

## Methods

### Study outcomes

The main objectives of the study are:

- To determine the prevalence of sarcopenia/presarcopenia in pregnant women with and without GDM;
- To identify pre-gestational and gestational factors that may be associated with the risk of developing or worsening existing sarcopenia;
- To determine the potential impact of sarcopenia/presarcopenia on pregnancy outcomes, including pregnancy complications, fetus growth by gestational age, type of delivery and delivery complications, newborn health, and metabolic status;
- To explore the relationship between sarcopenia, cognitive function and neuronal activity.

### Recruitment

Recruitment of the pregnant women will be completed in 8 months. Inclusion criteria will be Caucasian ethnicity, good comprehension of Italian or English language, willingness to participate to the study, and at least one among the following conditions: i) Age equal or higher than 35 years; ii) Body mass index (BMI) equal or higher than 25 kg/m^2^; iii) Glycemia between 5.6 and 6.9 mmol/L before or at the beginning of pregnancy; iv) Fetal macrosomia in a previous pregnancy; v) GDM in a previous pregnancy; vi) First-degree family history of diabetes.

Main exclusion criteria (in addition to unmatching of the inclusion criteria) will be twin pregnancy, presence of any already known disease/disorder possibly affecting muscle mass or function, neurological or psychiatric diseases, and already known diabetes (e.g., type 1 or type 2 diabetes). The main exit criterion will be the withdrawal of consent.

During the recruitment phase, we expect to perform at least 500 oral glucose tolerance tests (OGTT) for GDM screening. Based on our previous data, GDM prevalence is close to 25% of the screened women. Therefore, we expect to diagnose around 120 GDM women. Based on the inclusion and exclusion criteria and willingness to provide informed consent, we hypothesize 15-20% exclusion rate, leaving us with the potential for recruiting at least 100 GDM women. An equal number of non-GDM women (matched by age and BMI) will be recruited as well, thus for a total of at least 200 participants. The study will comply with the standards of good practice for medical research as defined in the Declaration of Helsinki. Written informed consent to study participation will be obtained for all recruited women. The study has already received approval by the local Ethics Committees (University of Pisa and Medical University of Vienna). The study protocol has also been registered at ClinicalTrials.gov.

### Normal pregnancy and gestational diabetes examinations

Before the scheduled date for the screening OGTT (7-10 days before), all women will be preliminarily contacted for careful description of the study, so that they will be in the position to provide written informed consent in the occasion of visit V1. In that occasion, recruited women will undergo 75g OGTT after an overnight fasting. High-risk women for GDM (previous GDM, and/or BMI equal/higher than 30 kg/m^2^, and/or glycemia between 5.6-6.9 mmol/L before or at the beginning of pregnancy) will undergo OGTT between week 16 and 18. All other women will undergo the OGTT at week 24-28. High-risk women, with normal OGTT at week 16-18, will repeat the test again at week 24-28. GDM diagnosis will be based on the criteria of the International Association of Diabetes and Pregnancy Study Group [20]. Blood samples for GDM diagnosis will be collected at zero (i.e., at fasting, immediately before glucose ingestion), 30, 60 and 120 min following glucose ingestion for measurement of plasma glucose, insulin and C-peptide levels. Before the OGTT, relevant medical and personal history (obstetric history, pre-pregnancy body weight, smoking, dietary and physical activity habits, education level, etc.) will be recorded along with measurement of BMI, blood pressure and heart rate.

At week 28-32, all participants will undergo hemochromocytometric test (visit V2). In that occasion, we will also measure glycated hemoglobin and adipokines (leptin, adiponectin, irisin), relevant for the relationships between sarcopenia and inflammation (21). The blood aliquot has been determined and deemed acceptable for pregnant women. Moreover, as for clinical routine, all women will undergo a further blood (and urine) test at week 35-37 (visit V3). In that occasion, if needed, we will have opportunity to repeat the test of one or more of the variables required for the study. It is also worth noting that at our Centers some additional visits/examinations are routinely performed for pregnant women, and particularly for GDM, including, among others, obstetric echography and cardiotocography. Though these further visits/examinations currently do not appear of major relevance for our study aims, related data will be available in the case that we identify any potential usefulness for the study.

### Muscle mass, strength, and physical performance (sarcopenia-related parameters)

Examinations related to muscle mass and strength, and to the degree of physical performance, will be accomplished in the third trimester of pregnancy, as this is the time the body composition undergoes main changes. Thus, since one of the main aims of the project is to determine muscle mass, strength and physical performance, the third trimester appears the most appropriate period for assessment of sarcopenia-related parameters. Precisely, muscle/physical performance tests will be performed at week 28-32. At the time being, we hypothesize performing these evaluations in the occasion of visit V2. However, if this may be of excessive burden to the pregnant women, it will be planned on a separate day, still between week 28-32.

Diagnosis of sarcopenia/presarcopenia will be based on the 2010 criteria of EWGSOP (European Working Group on Sarcopenia in Older People) [3]. For diagnosis of presarcopenia, simple low muscle mass is required. For sarcopenia, in addition to low muscle mass, low muscle strength and/or low physical performance need to be determined, and in case all abnormalities are present sarcopenia is defined as “severe”. Different techniques for sarcopenia/presarcopenia testing are indicated by EWGSOP. In our case, it is particularly important to opt for simple, non-invasive and not time-consuming techniques. Therefore, muscle mass will be tested by bioelectrical impedance analysis (BIA), muscle strength by handgrip strength test, and physical performance by the usual gait speed test [3]. In more detail, BIA will be performed by a professional body composition analyzer (with specific indication for use during pregnancy). This type of medical instrument can estimate skeletal muscle mass, and other body parameters, such as lean and fat body mass, intracellular and extracellular body water, dry lean mass (i.e., protein and mineral body content). Of relevance, BIA has been already used in several studies involving pregnant women, as summarized in a recent review [21]. Handgrip strength will be assessed by a professional hydraulic dynamometer. The usual gait speed test will be performed by asking the patient to walk at natural speed over a uniform path of prescribed length (typically, 6-10 m). The time required for walking that distance will be recorded by a chronometer.

Studies in sarcopenia/presarcopenia have suggested somehow different cut-off values for the parameters derived by the indicated tests. For the BIA-derived skeletal muscle mass, we will refer to the cut-off reported by Chien et al. [22], which suggests normalization of skeletal muscle mass (SM) by height squared (that is, skeletal muscle mass index, SMI, equal to SM/height^2^). In women, low SMI is assumed for SMI under 6.42 kg/m^2^. We have opted for this cut-off, compared to some slightly different ones [23, 24], because the former has been suggested by studying young rather than elderly people (though the study also included elderly people, analyzed separately [22]). Thus, for our study, the indicated cut-off appears more appropriate than others, in relation to the typical age of pregnant women. Cut-off for muscle mass strength, as derived by the handgrip test, is suggested by Lauretani et al. [25]. In women, low muscle strength is defined as handgrip strength below 20 kg. However, we will also consider cut-off values specific for different BMI intervals (28). Cut-off for gait speed is suggested again by Lauretani et al. [25] (low speed: below 0.8 m/s). Finally, we will assess the effect on sarcopenia/presarcopenia prevalence in our population when considering some cut-off values slightly updated, as reported in the 2019 guidelines of EWGSOP (named as EWGSOP2) [4].

### Cognitive function and lifestyle questionnaires

Cognitive impairment has been demonstrated in sarcopenia, especially related to low muscle strength [2]. This may be due to the direct cross-talk between muscle and brain, mediated by exercise-induced myokines release (also named as “exerkines”). We will measure cognitive function by one/two questionnaires, likely the Montreal Cognitive Assessment (MoCA) [26] and/or the Trail Making Test (TMT) [27]. All women will be asked to fill up the questionnaire(s) immediately after the sarcopenia-related tests.

Information on lifestyle will also be useful to complement the sarcopenia-related parameters as described above. Thus, in the same occasion of the sarcopenia and cognitive function tests, a questionnaire on nutritional habits and physical activity will be given to all women, asking them to fill it up and send it back within one week (by email or ordinary mail). Of note, nutritional counseling will be provided for the whole project duration.

### Functional near-infrared spectroscopy

On a voluntary basis, women will be also offered to undergo functional near-infrared spectroscopy (fNIRS) examination. Women agreeing to this option will perform fNIRS test at visit V2, during the handgrip test. The rationale for this examination is the fact that compromised nervous system function has been suggested as one of the important contributors to sarcopenia [28]. In fact, dopaminergic downregulation, inadequate motor programming and motor coordination impairment can lead to decline of supraspinal drive. In addition, motor unit reorganization and inflammatory changes in motor neuron can decrease conduction velocity and amplitude of compound muscle action potential. Furthermore, neuromuscular junctions remodeling may contribute to neuromuscular impairment [28]. The fNIRS tests will be performed by using OxyMon, plus fNIRS/EEG (electroencephalography) headcap (Artinis Medical Systems, The Netherlands). Women will wear the headcap, which allows detecting fNIRS (and optionally EEG) signals from various cortex areas by special electrodes (“optodes”). The headcap is connected to the OxyMon system that registers the signals and performs appropriate signal processing. The system measures oxy-, deoxy-, and total hemoglobin concentration changes. For higher reliability, women will perform the handgrip test twice, with prescribed time interval for resting. The system will register the fNIRS signals during the whole test session, made of the two handgrip tests and resting time in between. We will then analyze the data with both commercial tools and custom-made software for advanced analysis of signal variability (fractal dimension, detrended fluctuations, approximate entropy, etc.). Of note, exploitation of the handgrip test as motor task for fNIRS investigation in sarcopenia is consistent with what done in previous studies [29]. Also, fNIRS has been already used in pregnancy and specifically in GDM [30], thus indicating safety of this examination in those conditions.

### Data on pregnancy progression, delivery, newborn health status, and postpartum examinations

For all women included in the study we will collect all information regarding the usual evaluation of pregnancy, including obstetric echography, cardiotocography, ultrasound evaluation of the amniotic fluid, fetal growth. Moreover, the time and type of delivery will be recorded along with any complication. For the newborn, we will record sex, length, weight, and APGAR score (appearance, pulse, grimace, activity, respiration). Feeding information will also be collected.

At week 6-12 postpartum (visit V1POST), a diagnostic OGTT will be performed in women that had GDM, to screen for possible diabetes or prediabetes. In addition, for our specific study aims, a subgroup of women will be recalled for a follow-up visit (visit V2POST) at month 6 postpartum (±2 weeks). In that occasion, sarcopenia tests will be repeated, and further OGTT will be performed (similar to that at V1, i.e., 4 samples, and glucose, insulin, C-peptide measure). The other relevant blood tests will be repeated as well. Women will be also asked to fill up questionnaires again (plus fNIRS, optionally). We plan to recall about 50% of study participants, with criteria that will be precisely defined based on the baseline findings (likewise, women already sarcopenic/presarcopenic at baseline, and/or with abnormal values of sarcopenia-related parameters, e.g., in worst quartile for one or more of such parameters).

### Data analyses

Collected data will undergo quality control, based on visual inspection of data distributions, and on checks through automatic procedures that identify abnormalities, both in terms of absolute variables value and/or relative to other variables value, considering the specific population under analysis (pregnant and GDM women, in this case).

Moreover, by mathematical modelling of the OGTT data we will calculate insulin sensitivity, insulin secretion, pancreatic beta-cell function and insulin clearance. Insulin sensitivity deterioration (i.e., insulin resistance) is a typical trait of both sarcopenia and GDM (and partly of normal pregnancy as well). Thus, we will analyze this aspect in details, using different insulin sensitivity/resistance indices. Indeed, it is known that OGTT-based indices are surrogate markers of the “reference” insulin sensitivity as determined by clamp technique [31], which is however hardly feasible in the clinical routine. The validated OGTT-derived indices are typically good markers of the clamp-derived index, but on the other hand it is known that some differences may be shown among the different OGTT indices, especially in specific conditions [32]. Sarcopenia and GDM is certainly a peculiar “combined” condition that may determine unreliability of some indices. If all (or the majority) of indices will provide consistent information, this would be clue of their reliability. In contrast, if indices will provide conflicting results, this would call for the need of new, “specialized” indices. In this case, appropriate modelling and statistical analyses will be performed to derive a new index/method, by also exploiting the BIA data. Indices that will be computed are our recently developed PREDIM [33], and the more traditional OGIS [34], ISIcomp [35], MCRest [36], SIoral [37] (and, possibly, some others, such as CLIX [38]).

Insulin secretion and beta-cell function will be assessed by mathematical modelling that allows calculation of different aspects of the beta-cell behaviour [39, 40]. Briefly, the main beta-cell function parameters that will be derived from the model are: i) Beta-cell glucose sensitivity (the mean value of the dose-response function, describing the dependence of insulin secretion on absolute glucose concentration); ii) Rate sensitivity (proportionality constant of the derivative component of the secretion, representing the dynamic dependence of secretion on the rate of change of glucose); iii) Ratio of the potentiation function at 120 to that at zero minutes (explaining the sustained insulin secretion levels that are typically seen at the end of an OGTT in healthy subjects, when glucose has already returned to basal). Another interesting parameter is insulin secretion at reference glucose value. Insulin clearance will be derived by insulin secretion and plasma insulin levels. Furthermore, on the basis of the clinical variables that we will measure in addition to the OGTT, we will possibly compute other indices of potential interest in glucose metabolism, such as the triglycerides-glucose index [41], and the SPISE index [42].

### Statistical analyses

Data distributions will be assessed by graphical tests, and appropriate actions will be taken in case of skewed distributions (such as use of non-parametric tests, or log-transformation). Differences in the prevalence of sarcopenia in GDM vs. non GDM women will be assessed with Pearson’s Chi-squared test. Based on the OGTT and sarcopenia tests during pregnancy, participants will be stratified in two groups (GDM and nonGDM) and four subgroups: SARCO-GDM, nonSARCO-GDM, SARCO-nonGDM, nonSARCO-nonGDM (where “SARCO” indicates presence of sarcopenia (or presarcopenia), and “nonSARCO” indicates absence of sarcopenia/presarcopenia). We will test for possible differences among the glucometabolic parameters (such as insulin sensitivity/resistance), the sarcopenia-related parameters, and the other collected variables, among the indicated subgroups by Analysis of Variance (ANOVA), whereby Tukey’s post-hoc test will be used to achieve a 95% coverage probability. Analyses will be also performed in groups/subgroups appropriately pooled.

Moreover, correlation analyses (such as Spearman’s rho) will be used to assess possible associations among the sarcopenia-related parameters and the glucometabolic parameters, as well as the other collected variables (again, in groups/subgroups separated and/or pooled). Of note, we will perform both univariable and multivariable regression analyses, possibly driven by specific variable selection techniques, such as stepwise-regression analysis (or more advance shrinkage regression).

With the follow-up data, it will be possible to identify baseline predictors of sarcopenia/presarcopenia at later time (though in a relatively short period, due to project duration). Predictive accuracy will be tested by receiver operating characteristic (ROC) curves.

Furthermore, we will analyze the time trend of the sarcopenia-related parameters (muscle mass and strength, and physical performance) by appropriate methods such as linear mixed effects models.

Machine learning techniques will be also applied for robust regression and classification analyses. Statistical analyses will be performed with R and contributed packages. A two-sided type 1 error rate of 0.05 will be considered statistically significant. However, all p-values will be interpreted in an explorative manner in order to generate new hypotheses.

## Discussion

The study presented in this article will provide information on sarcopenia and presarcopenia prevalence in GDM and, possibly, in pregnancy uncomplicated by dysglycemia. Furthermore, the study will provide knowledge on the main glucometabolic factors associated to sarcopenia or presarcopenia (or, at any rate, associated to the possible worsening of the physiological variables on which sarcopenia/presarcopenia diagnosis is based). In addition, we will analyze factors other than the glucometabolic ones that may show possible association with sarcopenia (for instance, inflammatory factors). The baseline careful characterization of the study population coupled with the planned follow-up will also set the basis for identifying predicting factors. The identification of such factors will be of major importance as it may represent an initial guidance for treatments that may prevent sarcopenia in GDM/pregnant women. This will become of even greater interest to the extent that sarcopenia/presarcopenia may show to have an impact on progression and outcomes of pregnancy, in particular in women with GDM.

Notably, when sarcopenia develops, spontaneous remission has been rarely observed. Yet, several studies have shown that sarcopenia can be mitigated or even reverted by pharmacological intervention [43–45], or proper physical exercise, such as “resistance-type exercise” training [10]. Thus, with appropriate intervention possible GDM-induced sarcopenia/presarcopenia may be mitigated/reverted as well, and, likely, even prevented.

Another achievement of the study will be the possible identification of sarcopenia/presarcopenia as further risk factor for GDM (in addition to those already known as GDM risk factors [46–48]).

Although the focus of the study is to investigate how GDM may lead to sarcopenia, it is possible that, on the other side, sarcopenia contributes to GDM. This has in fact been suggested in the case of T2D, where the relationship between sarcopenia and T2D has been defined as “bidirectional” [49].

Another novel aspect included in this project is the evaluation of cognitive and neuronal function. In fact, much emphasis is currently paid on what has been suggested to be a new diabetic “complication” [50], but little is known about it in GDM. Our investigation is of relevance because it exploits recent information linking sarcopenia and cognitive function, combining functional tests with direct neuronal exploration with a state-of-the-art approach. Due to its nature and duration, the project cannot answer all questions on this aspect, but it can be the basis for further scientific investigations.

## Declarations

### Ethics approval and consent to participate

The protocol has been approved by the Comitato Etico Regione Toscana - Area Vasta Nord Ovest (CEAVNO) on the 25/07/2024, ID 25652, and by the Local Ethics Committee of the Medical University of Vienna on the 17/06/2024, n. 1458/2024. All participants to the study will be asked to provide written informed consent to participate in the study.

### Consent for publication

Not applicable.

### Availability of data and materials

Not applicable.

### Authors’ contributions

AD, CG and AT conceived and designed the study protocol. GD participated in its design. AT drafted the manuscript. AD, CG, GD revised the manuscript. All authors gave the final approval of the version to be submitted.

## Data Availability

Not applicable (study protocol)

## List of abbreviations

T2D: type 2 diabetes
GDM: gestational diabetes mellitus
BMI: body mass index
OGTT: oral glucose tolerance test
EWGSOP: European Working Group on Sarcopenia in Older People
BIA: bioelectrical impedance analysis
SM: skeletal muscle mass
SMI: skeletal muscle mass index
MoCA: Montreal Cognitive Assessment
TMT: Trail Making Test
fNIRS: functional near-infrared spectroscopy
EEG: electroencephalography
APGAR: appearance, pulse, grimace, activity, respiration
SARCO: sarcopenia
nonSARCO: no sarcopenia
nonGDM: no GDM
ROC: receiver operating characteristic

